# Vitamin D Deficiency among Industrial Workers in Cikarang, Indonesia: Prevalence, Occupational Determinants, and Health Implications

**DOI:** 10.64898/2026.05.05.26351787

**Authors:** Theresia Santi, Todia Pediatama Setiabudiawan, Jenifer Kiem Aviani, Muhammad Subhan Alfaqih, Cornelia Jaqualina, Angelie Firstania Rizqika Nanova, Khairunnisa Azkiya Putri Saila, Budi Setiabudiawan

## Abstract

**Objective:** To assess the prevalence, determinants, and health associations of vitamin D deficiency among workers in Cikarang, Indonesia.

**Methods:** Cross-sectional study of 107 workers. Serum 25(OH)D was measured by ECLIA; deficiency was <20 ng/mL. Data included demographics, occupation, sun exposure, diet, anthropometry, laboratory parameters, and self-reported health. Analyses used t-test/Mann–Whitney U, chi-square, and multivariable regression.

**Results:** Deficiency was prevalent (57.9%; median 18.6 ng/mL). Males had higher levels (+7.60 ng/mL) and lower risk (OR 8.56), while younger age (<38.5 years) showed lower levels (−5.60 ng/mL) and higher risk (OR 4.73; all p<0.0001). Greater sun exposure increased levels, whereas sunscreen use and greater clothing coverage reduced them. Only sex and age remained significant.

**Conclusion:** Vitamin D deficiency is common, especially among female and younger workers, with modifiable sun-avoidance behaviors contributing.

**LEARNING OUTCOMES:**

- Summarize the prevalence and key determinants of vitamin D deficiency among industrial workers in equatorial Indonesia, contributing to limited occupational health data in this setting.
- Highlight the role of modifiable behaviors—particularly sun exposure and sun-avoidance practices—in influencing vitamin D status among workers.
- Emphasize the need for targeted workplace interventions, including safe sun exposure strategies and nutritional support, especially for high-risk groups such as female and younger workers.

## INTRODUCTION

Vitamin D is a fat-soluble prohormone essential for calcium and phosphate homeostasis and bone mineralization. Beyond skeletal functions, it modulates immune responses, cardiovascular function, and glucose metabolism.^1–6^ The primary source of vitamin D is cutaneous synthesis, whereby ultraviolet B (UVB) radiation converts 7-dehydrocholesterol in the skin to previtamin D_3_, while dietary intake contributes minimally.^7^ Vitamin D deficiency, defined as serum 25-hydroxyvitamin D [25(OH)D] <20 ng/mL (50 nmol/L), has been associated with increased susceptibility to infections,^1,3,8^ cardiovascular disease,^2^ metabolic disorders,^9–11^ and anemia.^12^

Despite abundant sunlight in equatorial regions, vitamin D deficiency remains prevalent worldwide, independent of latitude. A pooled analysis of over seven million participants reported widespread insufficiency, with more than three-quarters of individuals in Asia affected.^13^ In Indonesia, deficiency has been documented among pregnant women, children, and adolescents.^14–16^ Cultural practices, including limited skin exposure and habitual sun avoidance, may contribute to this paradox.^17,18^

Workers may experience additional occupational constraints on sun exposure, particularly indoor and shift workers. Systematic reviews report higher deficiency rates among healthcare workers and indoor occupations compared with outdoor workers,^19,20^ with ethnicity and sex further modifying risk.^21,22^ However, there is a critical lack of data from equatorial industrial work settings, where sunlight availability is high but exposure is behaviorally and occupationally restricted. In Indonesia, evidence on vitamin D status among active workers remains scarce, and no studies have comprehensively examined the interplay between occupational exposure, sun-avoidance behaviors, and biological factors in determining vitamin D status. Furthermore, the relationship between vitamin D and routine hematological parameters and health-effects in working populations has not been clearly established, limiting its translational relevance for workplace health screening.

Given these gaps, this study aimed to determine the prevalence and determinants of vitamin D deficiency among industrial workers in Indonesia and to evaluate its association with hematological parameters and health-related outcomes.

## METHODS

### Ethical Declaration

This study was reviewed and approved by the Institutional Review Board (IRB) of Bekasi Regional General Hospital under approval number KP.800.1.6/3666/RSUD/2025. Written informed consent was obtained from all participants prior to enrolment in the study.

### Study Design and Participants

This cross-sectional study assessed the prevalence and determinants of vitamin D deficiency among workers in Cikarang, West Java, Indonesia, between June and August 2025. Adults aged 18–60 years who were actively employed were included, while individuals with a history of cancer or autoimmune disease were excluded to reduce potential confounding effects on vitamin D metabolism.

Sample size was calculated using an estimated deficiency prevalence of 87% from a comparable population,^23^ with 10% precision, 5% significance level, and 80% power, yielding a minimum requirement of 100 participants.

Participants were recruited through workplace announcements and company outreach using convenience sampling, with efforts to include diverse occupational sectors to enhance representativeness.

### Data Collection

Data were collected through physical examinations, structured questionnaires, and laboratory analyses conducted by trained healthcare professionals. Height and weight were measured using standard procedures to calculate body mass index (BMI), categorized according to World Health Organization guidelines.^24^

Questionnaires captured demographic characteristics, occupational details, lifestyle factors (including sun exposure habits and supplement use), dietary intake, and self-reported health conditions. Sun exposure was assessed using a standardized instrument evaluating frequency, duration, timing, and protective behaviors.^25^ Dietary intake was measured using a food frequency questionnaire focusing on vitamin D–rich foods.

Venous blood samples were collected to determine serum 25-hydroxyvitamin D [25(OH)D] and albumin levels. Serum 25(OH)D was measured using electrochemiluminescence immunoassay (ECLIA; COBAS E411). Vitamin D deficiency was defined as 25(OH)D <20 ng/mL (50 nmol/L), and insufficiency as 20–29 ng/mL, and sufficiency as ≥ 30 ng/mL according to Endocrine Society guidelines.^5^ Due to the small number of sufficient cases, vitamin D status was recategorized as deficient (<20 ng/mL) and adequate (≥20 ng/mL). Routine hematology and biochemical parameters were analyzed using standard laboratory methods.

### Statistical Analysis

Statistical analyses were performed using Python 3.13 (statsmodels 0.14.6, scipy 1.17.0) and IBM SPSS Statistics version 20.0 (IBM Corp., Armonk, NY, USA), and figures were generated using GraphPad Prism version 10.1.2 (GraphPad Software, LLC, San Diego, CA, USA).

Bivariate analyses were conducted as an initial exploratory step to identify variables associated with vitamin D levels. Vitamin D was analyzed as both a continuous and categorical variable. Normally distributed data (Shapiro–Wilk p > 0.05) are presented as mean ± SD and compared using the independent t-test or one-way ANOVA; non-normally distributed data are presented as median (IQR) and analyzed using the Mann–Whitney U test (with Hodges–Lehmann median difference and 95% CI) or the Kruskal–Wallis test, as appropriate. Vitamin D status was categorized as deficiency (<20 ng/mL) or adequacy (≥20 ng/mL). Categorical variables were compared using the Chi-square or Fisher’s exact test.

Variables for the multivariable model were selected based on a combination of exploratory bivariate findings and biological plausibility, subject to the events-per-variable (EPV) constraint of ≥10.^26,27^ The primary analysis was multivariable linear regression with continuous serum 25(OH)D as the outcome variable, preserving information that would be lost through dichotomization.^28^ If the residuals deviated from normality (Shapiro–Wilk p < 0.05), heteroscedasticity-consistent standard errors (HC3) were computed to verify the robustness of inference. Variance inflation factors (VIF) assessed multicollinearity.

The manuscript was prepared in accordance with the Strengthening the Reporting of Observational Studies in Epidemiology (STROBE) guidelines for cross-sectional studies,^29^ and the STROBE checklist is provided in **Supplementary Material 1**.

## RESULTS

### Vitamin D Level and Status Stratified by Baseline Characteristics

A total of 107 workers were included in the study. The workers were predominantly young to middle-aged workforce (average age of 35 years-old (range 22 – 58), with balanced sex ratio (50.5% female, 49.5% male). Most participants had a university-level education (57.9%). Half of the participants were office workers (50.5%), the majority worked indoors (88.8%), and 15.9% reported night-shift work (**Table 1**). The median serum 25(OH)D level was 18.6 ng/mL (range 8.1–55.1), with 57.9% classified as deficient.

Vitamin D levels differed significantly by sex, age, education, profession, and working place (**Figure 1**). Males had higher levels than females (median difference 7.60 ng/mL; 95% CI 5.40–9.70; p < 0.0001), with greater adequacy (77.8% vs. 22.7%) (**Figure 1A** and **Figure 1B**). Female workers had significantly higher odds of vitamin D deficiency than males (OR 8.556; 95% CI 3.508–20.863; p < 0.0001). Receiver operating characteristic (ROC) curve analysis was performed to determine the optimal age cut-off for vitamin D adequacy, using Youden’s Index to identify the point maximizing sensitivity and specificity. The cut-off value was 38.5 years. Workers aged <38.5 years had significantly lower serum 25(OH)D levels (median difference −5.60 ng/mL; 95% CI −8.70 to −3.00; p < 0.0001) (**Figure 1C**) and a higher prevalence of deficiency compared with those aged ≥38.5 years (74.2% vs. 25.8%) with odds ratio of 4.73 (95% CI 2.07 – 10.85; p <0.0001) (**Figure 1D**).

**Figure 1.**
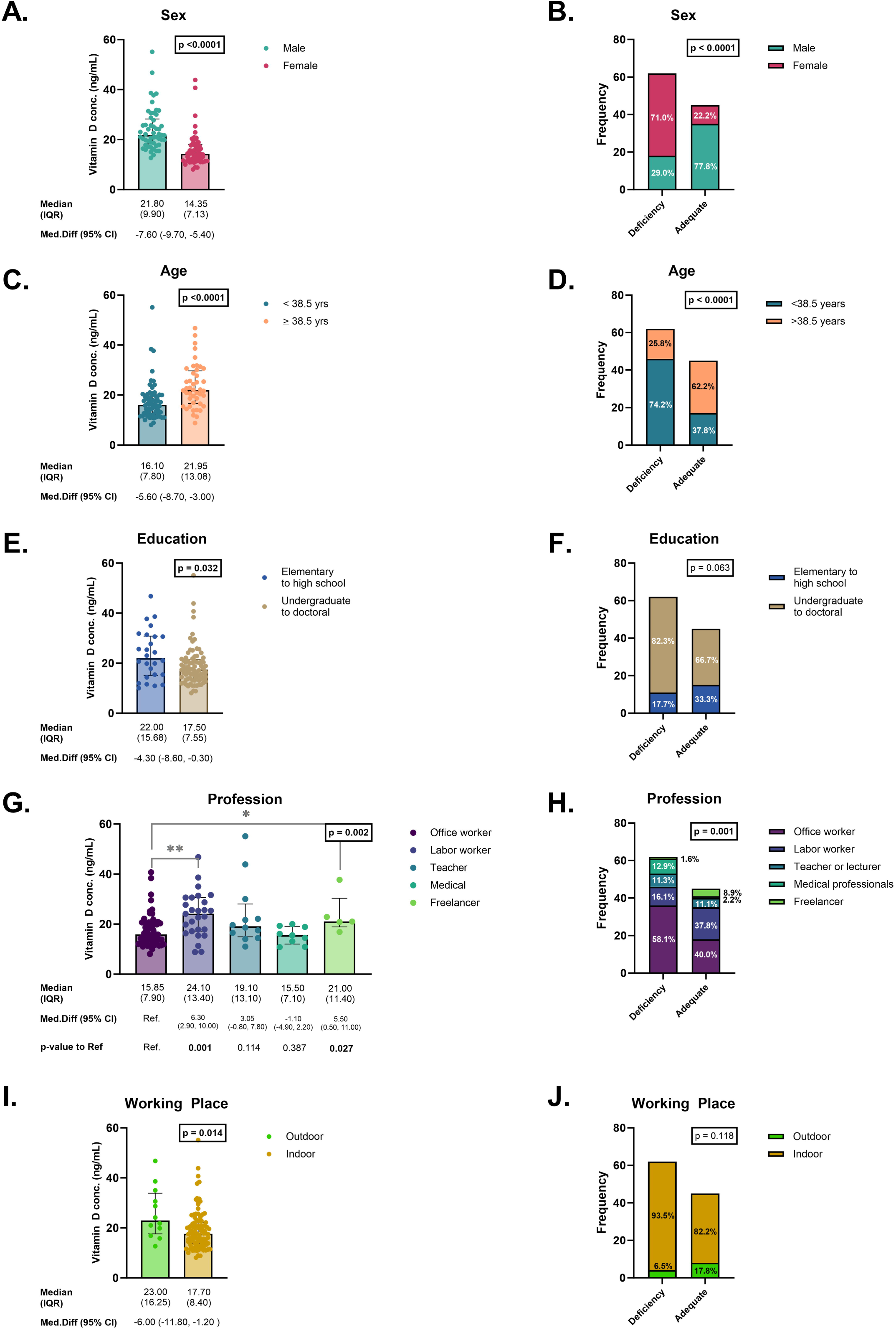
Serum vitamin D [25(OH)D] Levels and Deficiency Status Stratified by Baseline Characteristics. Data were presented as bar charts displaying median and interquartile range (IQR) for continuous serum 25(OH)D levels, with median differences and corresponding 95% confidence intervals shown below the graphs. Asterisks denote statistical significance between groups as follows: * = p <0.050, ** = p <0.010. Dichotomous vitamin D status was presented as stacked bar charts, with frequencies and percentages of participants indicated within the bars. Boxes above the graphs indicate between groups total p-value. **A.** Serum 25(OH)D level stratified by sex **B.** Distribution of sex across vitamin D status (deficient vs. adequate) **C.** Serum 25(OH)D level stratified by age at 38.5 years cut-off **D.** Distribution of age (at 38.5 years cut-off) across vitamin D status (deficient vs. adequate) **E.** Serum 25(OH)D level stratified by level of education **F.** Distribution of level of education across vitamin D status (deficient vs. adequate) **G.** Serum 25(OH)D level stratified by professions **H.** Distribution of professions across vitamin D status (deficient vs. adequate) **I.** Serum 25(OH)D level stratified by indoor or outdoor working place **J.** Distribution of indoor or outdoor working place across vitamin D status (deficient vs. adequate)

Lower educational attainment was associated with higher vitamin D levels (median difference 4.30 ng/mL; 95% CI 0.30–8.60; p = 0.032), without differences in adequacy proportion (p = 0.063) (**Figure 1E** and **Figure 1F**).

Vitamin D concentrations varied across professions (p = 0.002), with office workers showing lower levels (**Figure 1G**). Vitamin D status also differed by profession (p = 0.001) (**Figure 1H**). Outdoor workers had higher vitamin D levels than indoor workers (median difference 6.00 ng/mL; 95% CI 1.20–11.80; p = 0.014) (**Figure 1I**), although adequacy proportions were similar (p = 0.118) (**Figure 1J**).

### Vitamin D Level and Status Stratified by Sun Exposure Factors

Sun exposure during indoor work, outdoor work, and recreational activities was assessed by frequency, duration, timing, body surface area exposed, and total exposure score. Indoor exposure showed limited associations; only very short exposure (<5 minutes) was linked to lower vitamin D levels (p = 0.035), without differences in adequacy proportion. Total indoor exposure score was not significant (**Supplementary Figure 1**).

Outdoor exposure demonstrated stronger associations. Higher exposure frequency (p = 0.030) (**Figure 2A**) and greater body surface area exposure (p = 0.041) (**Figure 2C**) were associated with higher vitamin D levels. Exposed body surface area was also associated with adequacy (p = 0.023) (**Figure 2D**), wherase exposure frequency was not (**Figure 2B**). Workers who exposed only their face and hands had a 3.23-fold higher risk of vitamin D deficiency compared with those exposing the face, arms, and legs (OR 3.23; 95% CI 1.33–7.81; p = 0.009). Duration and timing were not independently associated. Total outdoor exposure score was not significant (**Supplementary Figure 2**).

**Figure 2.**
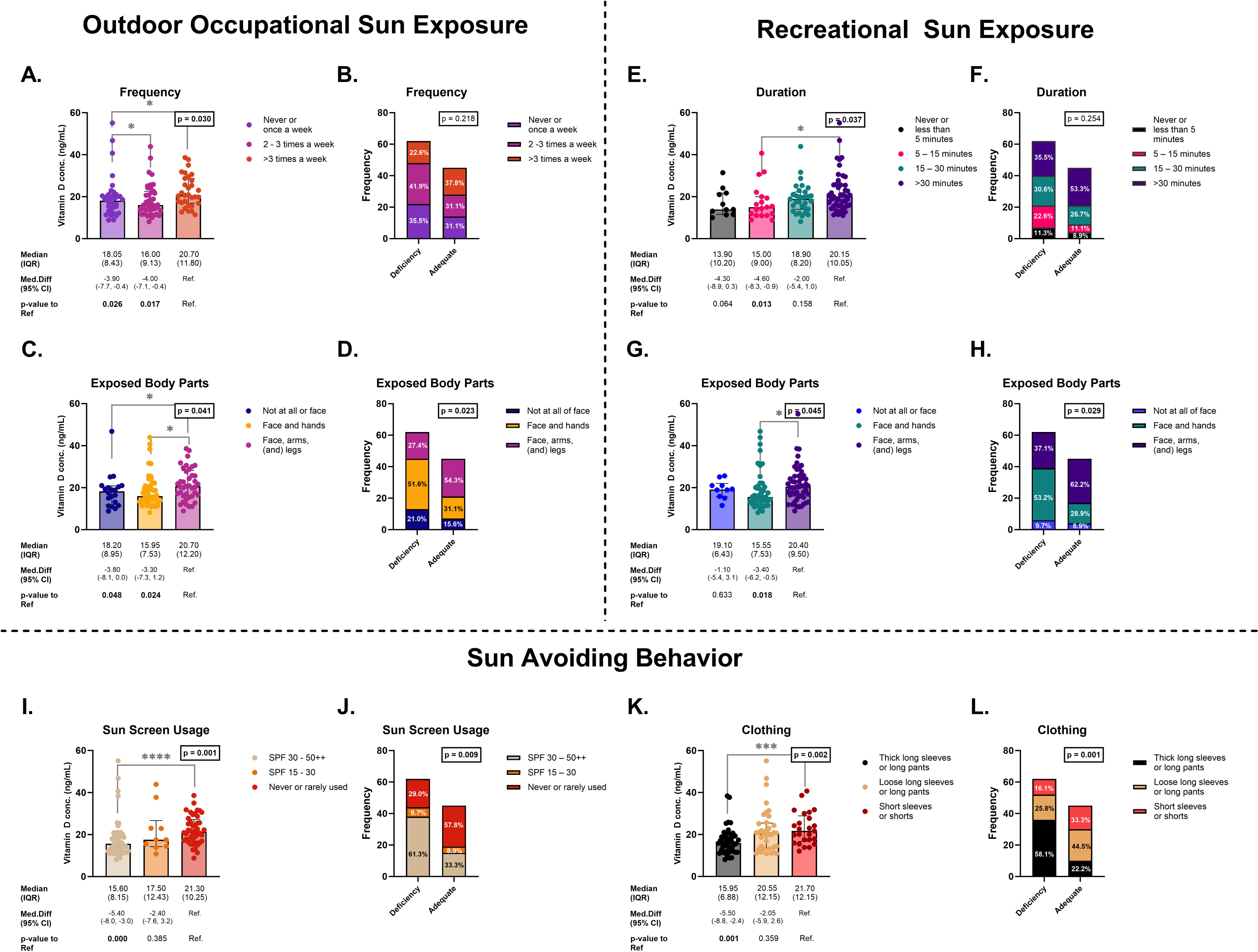
Serum vitamin D [25(OH)D] Levels and Deficiency Status Stratified by Significant Sun Exposure Factors. Data were presented as bar charts displaying median and interquartile range (IQR) for continuous serum 25(OH)D levels, with median differences and corresponding 95% confidence intervals shown below the graphs. Asterisks denote statistical significance between groups as follows: * = p <0.050, ** = p <0.010, *** = p <0.001, **** = p <0.0001. Dichotomous vitamin D status was presented as stacked bar charts, with frequencies and percentages of participants indicated within the bars. Boxes above the graphs indicate between groups total p-value. **A.** Serum 25(OH)D level stratified by outdoor occupational sun exposure frequencies **B.** Distribution of outdoor occupational sun exposure frequencies across vitamin D status (deficient vs. adequate) **C.** Serum 25(OH)D level stratified by outdoor occupational exposed body parts **D.** Distribution of outdoor occupational exposed body parts across vitamin D status (deficient vs. adequate) **E.** Serum 25(OH)D level stratified by recreational sun exposure durations **F.** Distribution of recreational sun exposure durations across vitamin D status (deficient vs. adequate) **G.** Serum 25(OH)D level stratified by recreational exposed body parts **H.** Distribution of recreational exposed body parts across vitamin D status (deficient vs. adequate) **I.** Serum 25(OH)D level stratified by Sun Protection Factor (SPF) levels of the sunscreen used **J.** Distribution of Sun Protection Factor (SPF) levels of the sunscreen used across vitamin D status (deficient vs. adequate) **K.** Serum 25(OH)D level stratified by clothing coverage **L.** Distribution of clothing coverage across vitamin D status (deficient vs. adequate)

For recreational sun exposure, longer duration was associated with higher serum 25(OH)D concentrations (p = 0.037) (**Figure 2E**), but was not associated with vitamin D adequacy (**Figure 2F**). In contrast, greater body surface area exposure was associated with both higher vitamin D levels (p = 0.045) (**Figure 2G**) and vitamin D adequacy (p = 0.029) (**Figure 2H**). Workers who exposed only their face and hands had a 3.09-fold higher risk of vitamin D deficiency compared with those exposing the face, arms, and legs (OR 3.09; 95% CI 1.33–7.20; p = 0.009). Frequency, timing, and total exposure score were not significant (**Supplementary Figure 3**). Overall, body surface area exposure during outdoor occupational and recreational activities was the most consistent sun-related determinant of vitamin D concentration.

Most participants had Fitzpatrick type III skin. Very fair to fair skin was associated with lower vitamin D levels (median difference −3.25 ng/mL; p = 0.042), though not adequacy proportion. Regarding sun avoiding behavior, staying in shade was not associated with vitamin D status (**Supplementary Figure 4**). Regular sunscreen use (p = 0.001) (**Figure 2I**) and greater clothing coverage (p = 0.002) (**Figure 2K**) were associated with lower vitamin D levels, and both were significantly associated with adequacy status (**Figure 2J** dan **Figure 2L**). Workers who wore thick long-sleeved clothing with long pants had a 5.40-fold higher risk of vitamin D deficiency compared with those wearing short sleeves and shorts (OR 5.40; 95% CI 1.86–15.64). The overall sun-avoidance score was lower among vitamin D–deficient workers (p = 0.001) (**Supplementary Figure 4**).

### Vitamin D Levels and Classification Stratified by Vitamin D Rich Food Consumption

Serum 25(OH)D concentrations did not differ according to supplementation status, and adequacy proportions were similar between supplement users and non-users (**Figure 3A** and **3B**). Intake of vitamin D–rich foods—including milk (**Figure 3C** and **3D**), seafood (**Figure 3E** and **3F**), meat (**Figure 3G** and **3H**), oil (**Figure 3I** and **3J**), and eggs (**Figure 3K** and **3L**)—was not associated with vitamin D levels or adequacy status. Likewise, the overall dietary intake score showed no significant relationship with serum 25(OH)D concentration (**Figure 3M**). These findings indicate that, in this population, supplementation and dietary intake were not independent determinants of vitamin D status

**Figure 3.**
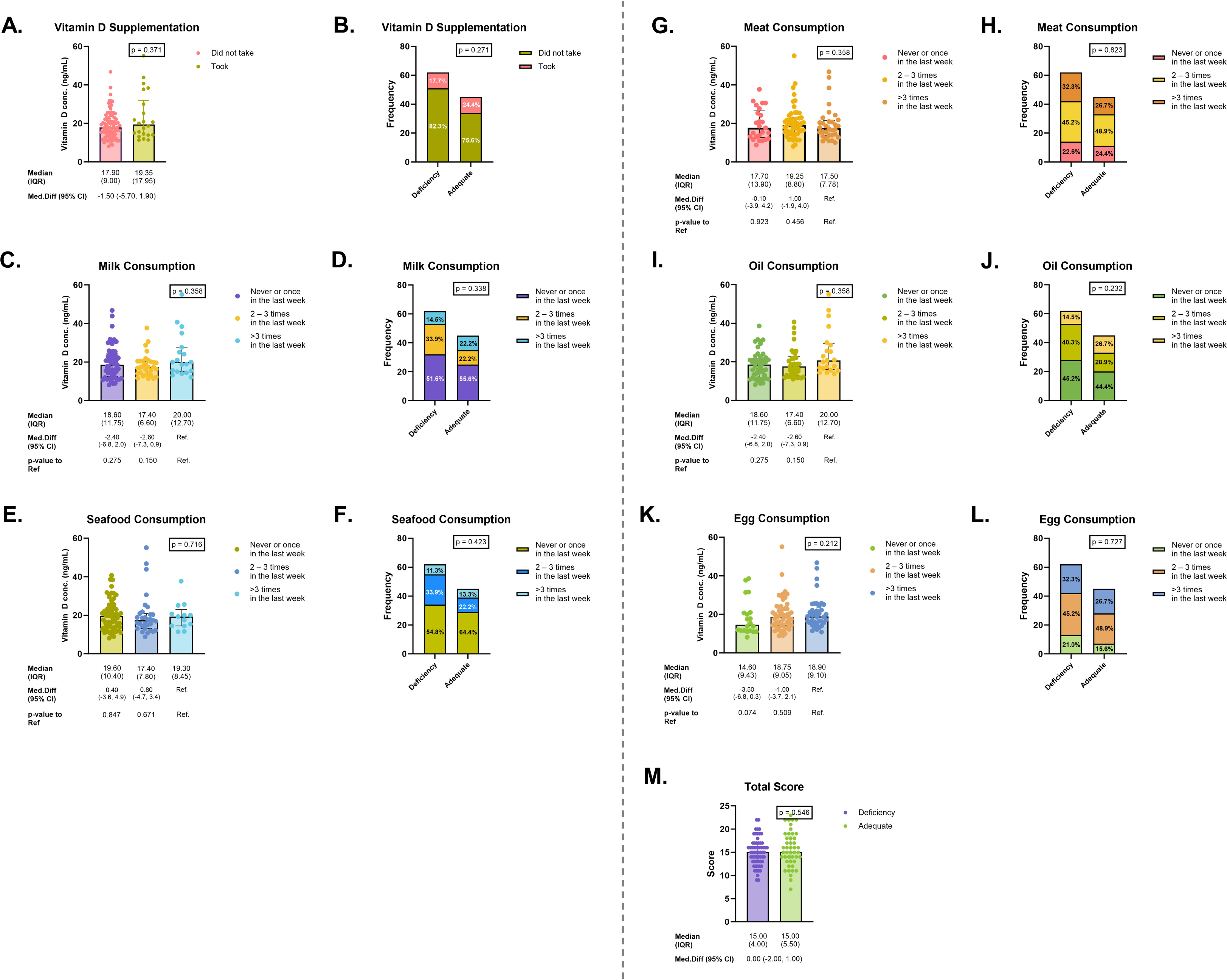
Serum vitamin D [25(OH)D] Levels and Deficiency Status Stratified by Consumption of Vitamin D-Rich Foods. Data were presented as bar charts displaying median and interquartile range (IQR) for continuous serum 25(OH)D levels, with median differences and corresponding 95% confidence intervals shown below the graphs. Dichotomous vitamin D status was presented as stacked bar charts, with frequencies and percentages of participants indicated within the bars. Boxes above the graphs indicate between groups total p-value. **A.** Serum 25(OH)D level stratified by vitamin D supplementation **B.** Distribution of vitamin D supplementation across vitamin D status (deficient vs. adequate) **C.** Serum 25(OH)D level stratified by milk consumption frequency **D.** Distribution of milk consumption frequencies across vitamin D status (deficient vs. adequate) **E.** Serum 25(OH)D level stratified by seafood consumption frequency **F.** Distribution of seafood consumption frequency across vitamin D status (deficient vs. adequate) **G.** Serum 25(OH)D level stratified by meat consumption **H.** Distribution of meat consumption frequency across vitamin D status (deficient vs. adequate) **I.** Serum 25(OH)D level stratified by oil consumption **J.** Distribution of oil consumption frequency across vitamin D status (deficient vs. adequate) **K.** Serum 25(OH)D level stratified by egg consumption **L.** Distribution of egg consumption frequency across vitamin D status (deficient vs. adequate) **M.** Total vitamin D-rich food consumption score stratified by vitamin D deficiency status

### Vitamin D Levels and Status Stratified by Body Anthropometrics

Serum 25(OH)D concentration was not associated with BMI category (p = 0.271) (**Figure 4A**), adequacy distribution (p = 0.146) (**Figure 4B**), or BMI values between deficiency and adequacy groups (p = 0.227) (**Figure 4C**), indicating no relationship between BMI and vitamin D status.

**Figure 4.**
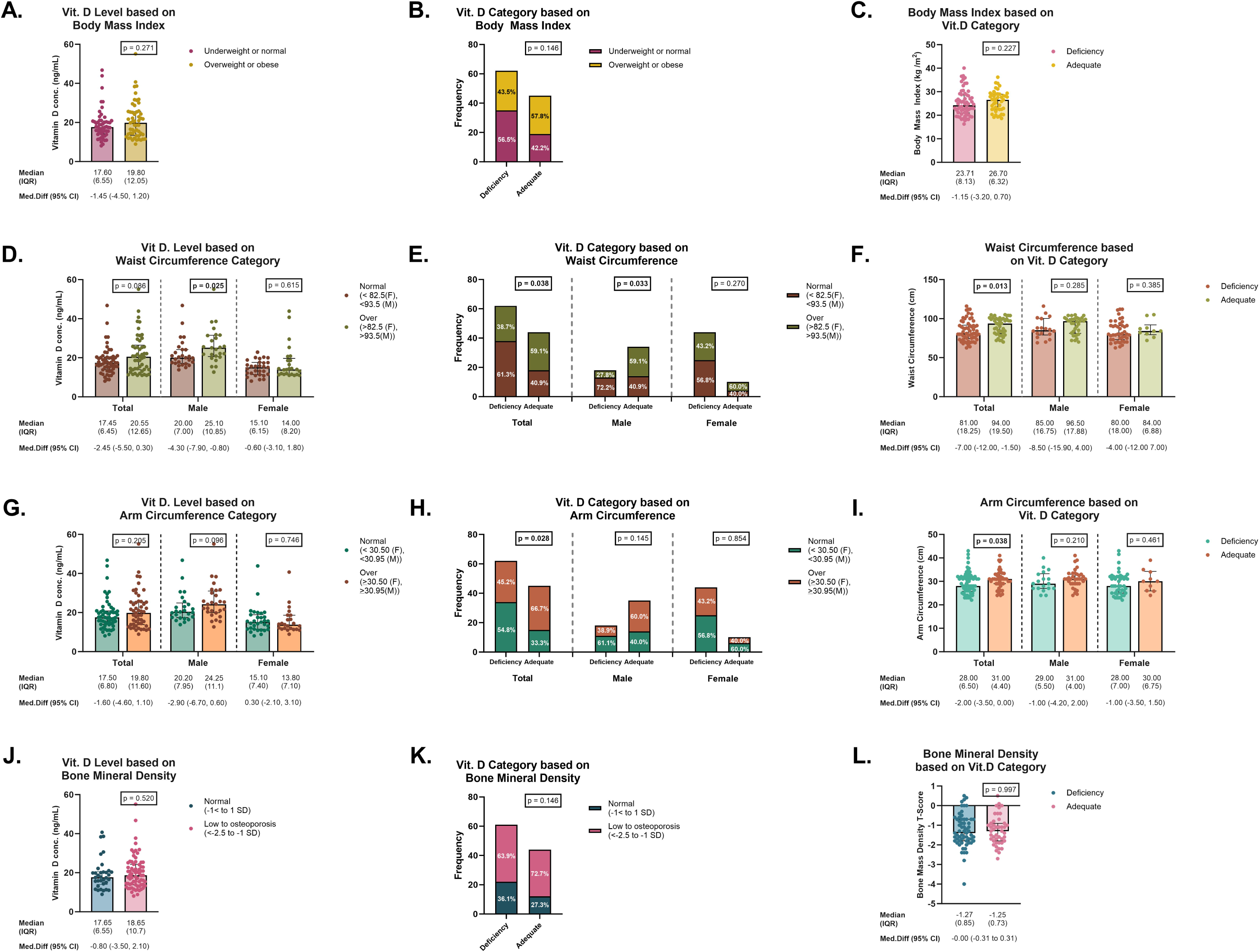
Serum vitamin D [25(OH)D] Levels and Deficiency Status Stratified by Body Anthropometrics. Data were presented as bar charts displaying median and interquartile range (IQR) for continuous serum 25(OH)D levels, with median differences and corresponding 95% confidence intervals shown below the graphs. Dichotomous vitamin D status was presented as stacked bar charts, with frequencies and percentages of participants indicated within the bars. Continuous anthropometric measurements were likewise presented as bar charts of median and IQR, stratified by vitamin D deficiency status. Boxes above the graphs indicate between groups total p-value. **A.** Serum 25(OH)D level stratified by body mass index category at 25 kg/m^2^ cut-off **B.** Distribution of body mass index category at 25 kg/m^2^ cut-off across vitamin D status (deficient vs. adequate) **C.** Body Mass Index stratified by vitamin D deficiency status **D.** Serum 25(OH)D levels stratified by waist circumference categories using cut-off values of 82.5 cm for females and 93.5 cm for males. The figure presents both the overall population and sex-stratified analyses. **E.** Distribution of waist circumference categories across vitamin D status (deficient vs. adequate). The figure presents both the overall population and sex-stratified analyses. **F.** Waist circumferences stratified by vitamin D deficiency status. The figure presents both the overall population and sex-stratified analyses. **G.** Serum 25(OH)D levels stratified by arm circumference categories using cut-off values of 30.50 cm for females and 30.95 cm for males. The figure presents both the overall population and sex-stratified analyses. **H.** Distribution of arm circumference categories across vitamin D status (deficient vs. adequate). The figure presents both the overall population and sex-stratified analyses. **I.** Arm circumferences stratified by vitamin D deficiency status. The figure presents both the overall population and sex-stratified analyses. **J.** Serum 25(OH)D level stratified by bone mineral density at ±1 SD cut-off **K.** Distribution of bone mineral density categories across vitamin D status (deficient vs. adequate). The figure presents both the overall population and sex-stratified analyses. **L.** Bone mineral density stratified by vitamin D deficiency status

Waist circumference categories were defined using ROC-derived cut-offs (Youden’s Index) with BMI as the reference distinguishing obese and non-obese individuals, yielding thresholds of 82.5 cm for women and 93.5 cm for men. Elevated waist circumference was not associated with higher vitamin D levels in the overall population (p = 0.086). Stratified analysis showed a significant association among males (p = 0.025), but not females (p = 0.615) (**Figure 4D**). Adequacy distribution differed by waist circumference in the total population (p = 0.038) and among males (p = 0.033) (**Figure 4E**). Adequate vitamin D status was associated with higher waist circumference in the overall population (p = 0.013). However, this association was not observed after stratification by sex (p = 0.285 in males and p = 0.385 in females), suggesting that the overall relationship may be influenced by sex-related distribution rather than an independent effect of vitamin D status (**Figure 4F**).

Arm circumference cut-offs (30.50 cm for women; 30.95 cm for men) were similarly derived by ROC analysis. Serum 25(OH)D concentrations did not differ significantly between individuals with normal and elevated arm circumference (**Figure 4G**). Although the distribution of vitamin D adequacy differed in the overall population (p = 0.028) (**Figure 4H**), no significant differences were observed after sex stratification for either continuous or categorical outcomes. Participants with adequate vitamin D status tended to have larger arm circumference (median difference 2.00 cm; 95% CI 0.00–3.50; p = 0.038) in the overall population; however, this association was not observed after stratification by sex (**Figure 4I**).

Bone mineral density (BMD) was not associated with vitamin D concentration or adequacy (all p > 0.05) (**Figure 4J** and **4K**) and no significant difference was found in BMD of individuals with vitamin D adequacy (**Figure 4L**). Overall, anthropometric and bone parameters were not consistently or independently associated with vitamin D status in this population.

### Multivariable Predictors of Vitamin D Concentrations

Based on the exploratory univariate findings and biological plausibility and with 45 vitamin D adequate participants (the smaller outcome group), the model was restricted to a maximum of four predictors. Sex was selected as the primary variable of interest, given its dominant association in bivariate analysis and well-documented sex differences in vitamin D status. Age was included as a potential confounder, given its significant bivariate association. Total sun exposure score was included as the principal modifiable environmental determinant of cutaneous vitamin D synthesis. Sun avoidance behavior score, a composite of sunscreen use, shade-seeking, and protective clothing, was selected because bivariate analyses suggested that several sex-associated behavioral variables (clothing type, sunscreen use, work location) lost significance after sex adjustment, indicating potential mediation through sun avoidance behavior.

In the primary linear regression model, male sex was the strongest predictor of serum 25(OH)D concentration (B = 8.72, 95% CI: 5.53–11.92, p < 0.001), indicating that men had on average 8.7 ng/mL higher vitamin D levels than women after adjustment for age, sun exposure, and sun avoidance behavior. Age was independently associated with higher vitamin D (B = 0.21 per year, 95% CI: 0.06–0.37, p = 0.008). Total sun exposure score was not significant (B = 0.08, p = 0.47). Sun avoidance behavior showed a borderline inverse association (B = −0.72, 95% CI: −1.46 to 0.01, p = 0.054). The model explained 28.4% of variance in vitamin D levels (adjusted R² = 0.256, F(4,102) = 10.13, p < 0.001, **Figure 5**).

**Figure 5.**
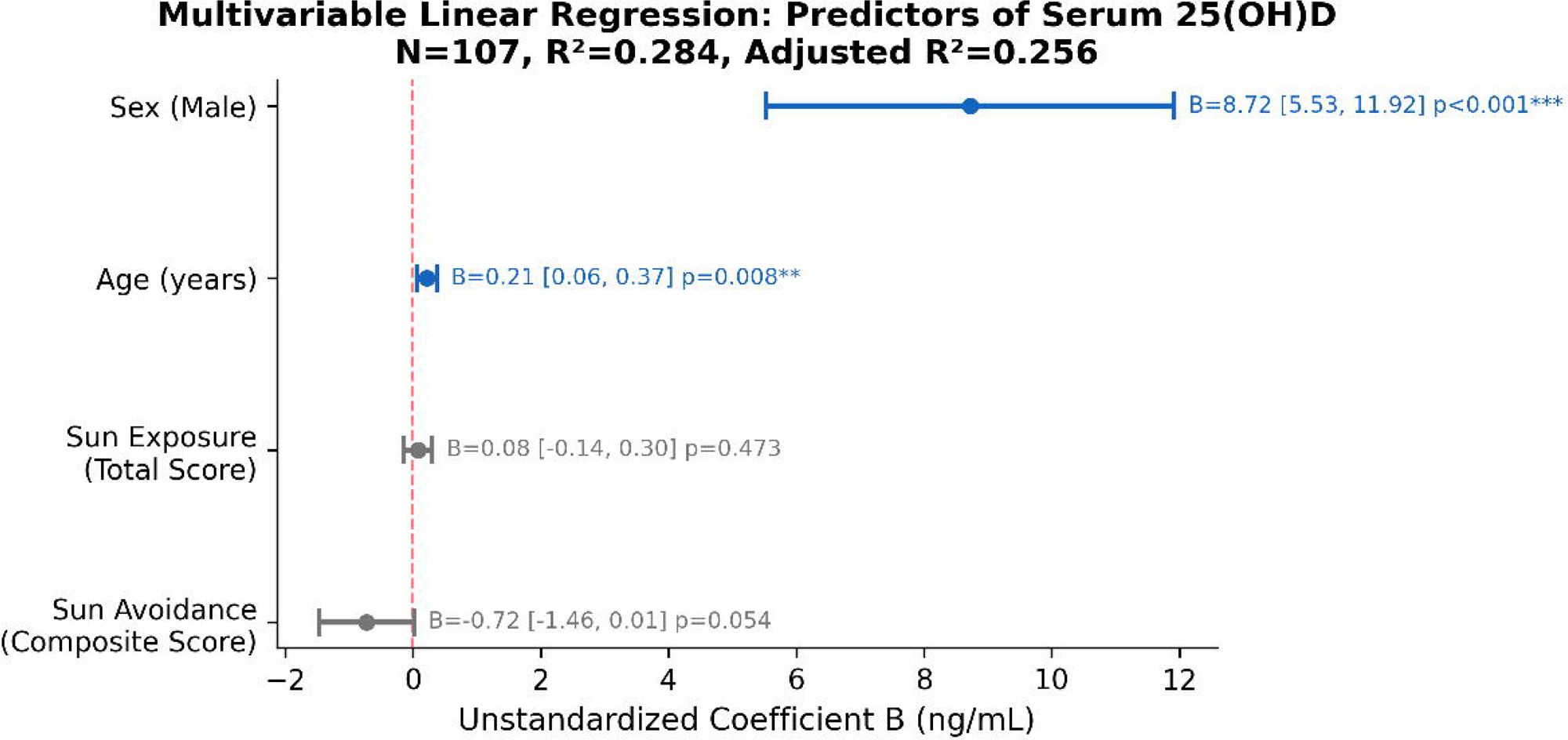
Forest plot of unstandardized beta coefficients from multivariable linear regression identifying the strongest predictors of serum vitamin D [25(OH)D] levels.

All VIFs were below 1.6, indicating no multicollinearity. With heteroscedasticity-robust standard errors (HC3), sex (p < 0.001) and age (p = 0.014) remained significant, while sun avoidance became non-significant (p = 0.24), suggesting its borderline association in the standard model was partially influenced by non-normal residual distribution.

### Association Between Vitamin D Level and Status with Laboratory Parameters

Serum 25(OH)D concentration showed modest but significant associations with selected hematological parameters. Participants with vitamin D adequacy had higher RBC counts (median difference 0.25 x 10^6^ / µL; 95% CI 0.02–0.43 x 10^6^ / µL; p = 0.033) (**Figure 6A**), hemoglobin levels (median difference 1.10 g / dL; 95% CI 0.50–1.70; g / dL p < 0.001 (**Figure 6D**), and hematocrit (median difference 2.90%; 95% CI 1.40–4.40%; p < 0.001) (**Figure 6G**), compared with those with deficiency.

**Figure 6.**
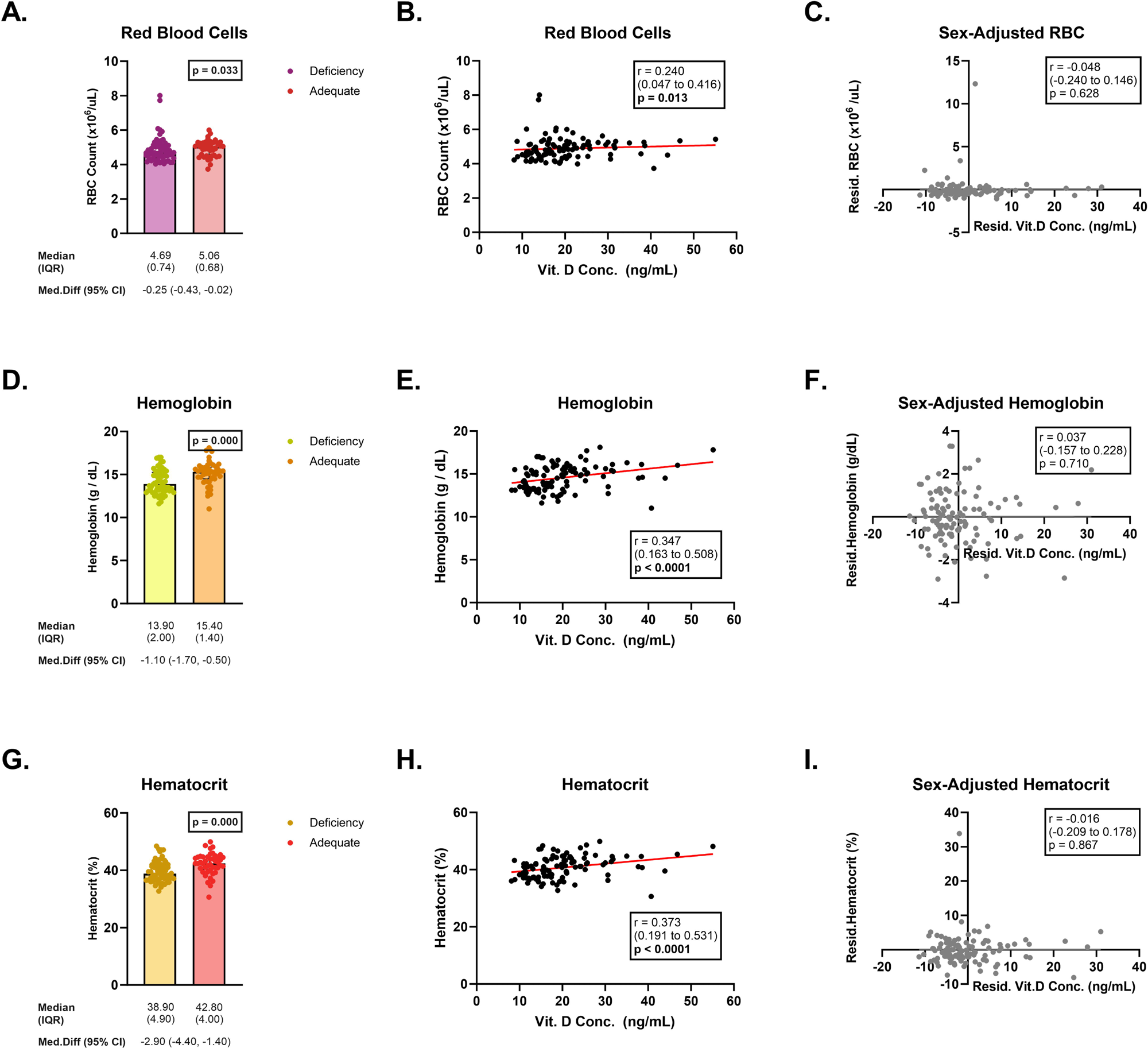
Association Between Serum vitamin D [25(OH)D] Levels and Deficiency Status with Significant Laboratory Parameters. Data were presented as bar charts displaying median and interquartile range (IQR) for continuous serum 25(OH)D levels, with median differences and corresponding 95% confidence intervals shown below the graphs. Associations between laboratory parameters and serum 25(OH)D levels were presented as scatter plots with fitted linear regression lines. Sex-adjusted analyses were displayed as scatter plots of residual serum 25(OH)D concentrations against residual laboratory parameters. Correlation coefficients and corresponding *p*-values are indicated within the plots. **A.** Red blood cells (RBC) Count stratified by vitamin D deficiency status **B.** Association between serum 25(OH)D concentration and RBC count **C.** Sex-adjusted association between serum 25(OH)D concentration and RBC count **D.** Haemoglobin level stratified by vitamin D deficiency status **E.** Association between serum 25(OH)D concentration and haemoglobin level **F.** Sex-adjusted association between serum 25(OH)D concentration and haemoglobin level **G.** Haematocrit level stratified by vitamin D deficiency status **H.** Association between serum 25(OH)D concentration and haematocrit level **I.** Sex-adjusted association between serum 25(OH)D concentration and haematocrit level

Unadjusted Spearman correlations demonstrated significant positive associations between serum 25(OH)D levels and RBC (r = 0.234; p = 0.013) (**Figure 6B**), hemoglobin (r = 0.347; p < 0.001) (**Figure 6E**), and hematocrit (r = 0.373; p < 0.001) (**Figure 6H**). However, after adjustment for sex, these associations were no longer significant: RBC (partial r = −0.048; p = 0.628) (**Figure 6C**), hemoglobin (partial r = 0.037; p = 0.710) (**Figure 6F**), and hematocrit (partial r = −0.016; p = 0.867) (**Figure 6I**). These findings indicate that the observed unadjusted correlations were confounded by sex, with no independent association between vitamin D and hematological parameters after adjustment.

No significant associations were found between vitamin D concentration and white blood cell count, platelet count, MCV, MCH, MCHC, RDW-CV, albumin, or random blood glucose levels (**Supplementary Figure 5**).

### Association Between Vitamin D Level and Status with Self-Reported Health Condition

Serum 25(OH)D concentration and adequacy were not significantly associated with self-reported respiratory (**Figure 7A** and **7B**), gastrointestinal (**Figure 7C** and **7D**), musculoskeletal (**Figure 7E** and **7F**), anxiety (**Figure 7G** and **7H**), sleep (**Figure 7I** and **Figure 7J**), or somatic symptoms (**Figure 7K** and **7L**). Although workers reporting frequent somatic disturbances had slightly higher vitamin D levels (median difference 2.90; 95% CI 0.20–5.60; p = 0.038), this association was not observed when vitamin D was analyzed categorically (p = 0.164).

**Figure 7.**
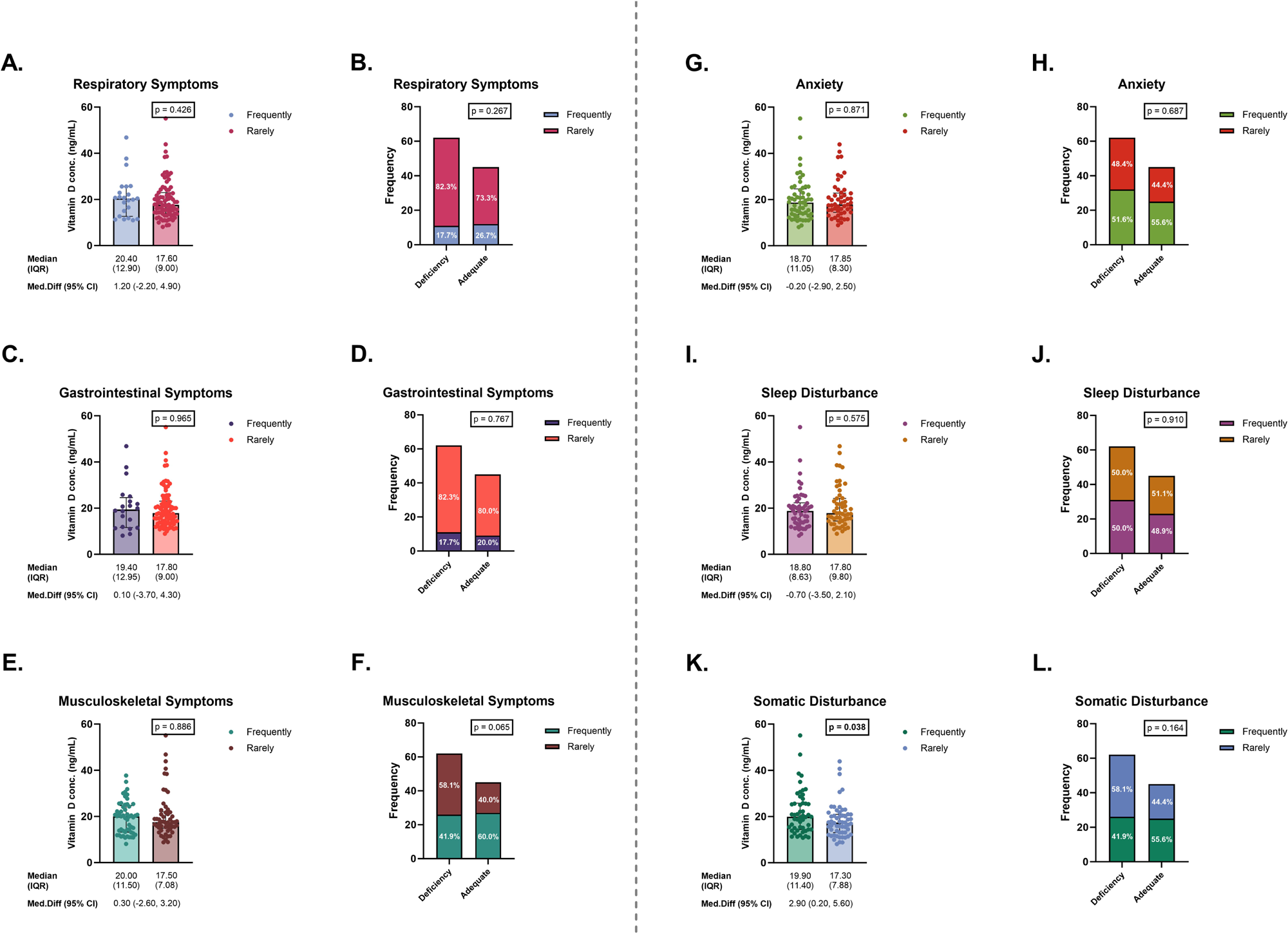
Serum vitamin D [25(OH)D] Levels and Deficiency Status Stratified by Self-Reported Health Conditions. Data were presented as bar charts displaying median and interquartile range (IQR) for continuous serum 25(OH)D levels, with median differences and corresponding 95% confidence intervals shown below the graphs. Dichotomous vitamin D status was presented as stacked bar charts, with frequencies and percentages of participants indicated within the bars. Boxes above the graphs indicate between groups total p-value. **A.** Serum 25(OH)D level stratified by self-reported respiratory symptoms **B.** Distribution of respiratory symptom frequency across vitamin D status (deficient vs. adequate) **C.** Serum 25(OH)D level stratified by self-reported gastrointestinal symptoms **D.** Distribution of gastrointestinal symptoms symptom frequency across vitamin D status (deficient vs. adequate) **E.** Serum 25(OH)D level stratified by self-reported musculoskeletal symptoms **F.** Distribution of musculoskeletal symptom frequency across vitamin D status (deficient vs. adequate) **G.** Serum 25(OH)D level stratified by self-reported anxiety **H.** Distribution of anxiety frequency across vitamin D status (deficient vs. adequate) **I.** Serum 25(OH)D level stratified by self-reported sleep disturbance **J.** Distribution of sleep disturbance frequency across vitamin D status (deficient vs. adequate) **K.** Serum 25(OH)D level stratified by self-reported somatic disturbance **L.** Distribution of somatic disturbance frequency across vitamin D status (deficient vs. adequate)

## DISCUSSION

Here we showed that vitamin D deficiency is highly prevalent among industrial workers in equatorial Indonesia, with 57.9% of 107 workers in Cikarang, West Java, falling below 20 ng/mL and 87% below the sufficient threshold of 30 ng/mL. The median 25(OH)D concentration was 18.6 ng/mL. In a global pooled analysis of nearly 7.9 million participants, vitamin D deficiency was widespread across all world regions, including tropical areas.^30^ Across Asia, 76.85% of individuals across heterogeneous populations fell below 30 ng/mL,^13^ broadly consistent with our 87%, though direct comparison is limited by differences in population composition and assay methods. In Indonesia specifically, deficiency has been reported in 63% of pregnant women^14^ and similarly high rates among children and adolescents.^15^

Among occupational cohorts, a systematic review of 71 studies encompassing over 53,000 workers found deficiency in 78% of indoor and 48% of outdoor workers,^19^ and shift workers show particularly high rates.^20^ Our findings extend this evidence to an Indonesian industrial workforce, a population that has received limited attention. In Cikarang, UVB availability is not the limiting factor: Indonesia’s equatorial position ensures favorable solar zenith angles year-round. What varies is behavior, specifically how much workers expose themselves to available sunlight.

The sex difference in vitamin D status was the central finding of this study. After adjustment, male workers had 25(OH)D concentrations approximately 7.1 ng/mL higher than female workers (adjusted B = 8.72, 95% CI: 5.53–11.92, p < 0.001). Unadjusted odds ratio showed female sex was associated with nearly twelve-fold higher odds of deficiency.

Occupational setting (indoor vs. outdoor) was associated with vitamin D status in bivariate analyses but lost significance after multivariable adjustment. This likely reflects the distribution of job types in this industrial setting, where many roles tend toward indoor or administrative work. Similar patterns have been reported among female indoor workers in Asian populations,^31^ suggesting that the observed occupational gradient may not represent an independent effect.

Clothing coverage and sun exposure—across both occupational and recreational settings—are key determinants of vitamin D status but lost significance after adjustment, suggesting a mediating role rather than independent effects. Although biological mechanisms (e.g., estrogen effects, vitamin D–binding protein variation, and *GC* gene polymorphisms) may contribute, the literature supports behavioral explanations. Wearing a hijab was the strongest predictor of deficiency in a Lebanese study,^32^ while exposed body surface area correlated with 25(OH)D in Indonesian women (r = 0.39).^33^ Similar findings across Saudi Arabia, India, and Malaysia indicate consistent effects of culturally driven sun avoidance.^31,34,35^ In our cohort, 64.8% of women wore hijabs, limiting UVB exposure.

Bivariate analysis showed that frequency and duration of sun exposure were significantly associated with 25(OH)D levels, consistent with studies from Metro Manila,^36^ Malay women,^37^ and Ecuador.^38^ However, sun exposure likely acts as a mediator in our study, as more women reported sun-avoidance behaviors and greater clothing coverage, both requiring longer exposure to achieve adequate synthesis.^33^ While 10–15 minutes of midday sun exposure (2–3 times/week) may suffice without full coverage,^5,17^ longer durations are likely needed in this context.

Sunscreen use and sun-avoidance behaviors further support this pattern. Although associated in bivariate analysis, sun-avoidance lost significance after adjustment, consistent with mediation. Sun avoidance remains common in Asian populations due to aesthetic concerns, with practices such as umbrella use, seeking shade,^39^ and long-sleeved clothing widely reported in Indonesia.^40^ Together, these findings highlight the dominant role of modifiable behavioral factors in shaping vitamin D status.

Age was positively associated with vitamin D levels in this population (B = 0.21 per year, p = 0.008). Our findings are consistent with previous studies showing higher serum 25(OH)D levels with increasing age among individuals with normal renal function (20–64 years).^41^ Similarly, a large study of 102,342 participants reported a higher prevalence of vitamin D deficiency in younger adults (18–29 years, 81.8%), which decreased with age (59.3% in those aged 50–65 years).^42^ This may reflect a healthy worker effect, whereby older workers who remain in industrial employment are inherently healthier, or seniority-related differences in occupational sun exposure. The narrow age range of working adults (predominantly 18–55 years) may also preclude detection of the age-related decline in cutaneous vitamin D synthesis that becomes prominent after age 65.^43,44^

Dietary vitamin D intake and supplementation showed no significant associations, consistent with the broader literature indicating that dietary sources contribute minimally to vitamin D status in populations with potential for cutaneous synthesis,^7,45^ and that supplementation use in this sample was too low and inconsistent to produce detectable effects.

Body mass index was not associated with 25(OH)D. In contrast, waist and arm circumference showed a paradoxical positive association with vitamin D adequacy in male workers, opposite to the inverse relationship expected from adipose sequestration.^46^ This finding should be interpreted cautiously given the small subsample and may reflect residual confounding. In this context, larger arm circumference likely reflects greater muscle mass and physically demanding or outdoor work, leading to increased UVB exposure. Waist circumference may similarly coexist with higher lean mass rather than purely adiposity, attenuating the expected inverse association. Lower engagement in sun-avoidance behaviors among men may further contribute. Overall, these anthropometric measures likely act as proxies for habitual sun exposure rather than independent determinants of vitamin D status.

Positive associations were initially observed between 25(OH)D and red blood cell count, hematocrit, and hemoglobin, consistent with the proposed role of vitamin D in erythropoiesis via effects on erythroid progenitor proliferation and hepcidin-mediated iron metabolism.^47–49^ However, males in this cohort exhibited both higher 25(OH)D levels and higher hematological parameters. After adjustment for sex, these associations were attenuated, indicating that the relationships were largely driven by sex differences rather than an independent effect of vitamin D.

No significant associations were observed between serum 25(OH)D and self-reported health conditions. Prior reviews, including RCTs, show that vitamin D supplementation reduces musculoskeletal symptoms and upper respiratory infections, but these were conducted in selected or clinical populations.^50–54^ In contrast, our cross-sectional study in a generally healthy workforce with low symptom prevalence, self-reported outcomes, and no intervention likely limited statistical power and attenuated differences, resulting in similar odds between vitamin D–deficient and adequate groups.

Our study has several limitations. First, the cross-sectional design precludes causal inference and does not establish temporality.^55^ Sun exposure was assessed by self-report, subject to recall bias and unable to capture UVB intensity or precise body surface area exposed, and we did not use objective measures such as personal UV dosimetry. Our questionnaire did cover multiple dimensions systematically, including frequency, duration, timing, body surface area, and sun protection practices. Dietary vitamin D was estimated from a food frequency questionnaire, which carries known measurement error for micronutrients with few food sources, though dietary sources contribute minimally to vitamin D status in equatorial populations, consistent with our null finding. Physical activity and smoking status were not measured, all of which may influence vitamin D metabolism or outdoor behavior.^56,57^ Further, a single time-point measurement may not reflect year-round status, though seasonal variation in 25(OH)D is attenuated at equatorial latitudes compared with temperate regions.^58^

Second, vitamin D status was assessed by electrochemiluminescence immunoassay (ECLIA) rather than LC-MS/MS, the reference method. ECLIA may introduce measurement error through cross-reactivity with inactive metabolites, but has established reliability for epidemiological classification and is widely used in population-based studies.^59^

Third, the sample size of 107 workers limits statistical power, particularly for within-sex sub-analyses where the number of vitamin D–adequate women was small (n = 10). The sample nonetheless exceeded the calculated minimum requirement and was sufficient to detect the central sex difference with strong statistical significance (p < 0.001).

Last, workers were recruited by convenience sampling from a single industrial region, limiting generalizability to other settings. This is, however, among the first studies to characterize vitamin D status in Indonesian industrial workers, filling a clear gap in the literature.

In conclusion, vitamin D deficiency was highly prevalent among industrial workers in equatorial Indonesia. Sex was the dominant predictor, with females having significantly lower 25(OH)D levels than males after multivariable adjustment. These findings support the need for proactive workplace-based interventions, including structured safe sun exposure programs (e.g., scheduled outdoor breaks), routine 25(OH)D screening for at-risk workers, and incorporation of vitamin D supplementation into occupational health services. Particular attention should be directed toward female workers, especially those with limited sun exposure due to cultural or religious practices, for whom supplementation may be the most effective strategy. Given the high burden observed, integrating vitamin D prevention into workplace health policies should be considered. Further longitudinal studies with objective exposure assessment are warranted to refine and optimize these interventions.

## Data Availability

The data supporting the findings of this study are not publicly available due to privacy and ethical restrictions but are available from the corresponding author upon reasonable request.

## FIGURE LEGENDS

**Supplementary Figure 1.** Serum vitamin D [25(OH)D] Levels and Deficiency Status Stratified by Indoor Occupational Sun Exposure

Data were presented as bar charts displaying median and interquartile range (IQR) for continuous serum 25(OH)D levels, with median differences and corresponding 95% confidence intervals shown below the graphs. Asterisks denote statistical significance between groups as follows: * = p <0.050, ** = p <0.010, *** = p <0.001, **** = p <0.0001. Dichotomous vitamin D status was presented as stacked bar charts, with frequencies and percentages of participants indicated within the bars. Boxes above the graphs indicate between groups total p-value.

**Supplementary Figure 2.** Serum vitamin D [25(OH)D] Levels and Deficiency Status Stratified by Outdoor Occupational Sun Exposure

Data were presented as bar charts displaying median and interquartile range (IQR) for continuous serum 25(OH)D levels, with median differences and corresponding 95% confidence intervals shown below the graphs. Asterisks denote statistical significance between groups as follows: * = p <0.050, ** = p <0.010, *** = p <0.001, **** = p <0.0001. Dichotomous vitamin D status was presented as stacked bar charts, with frequencies and percentages of participants indicated within the bars. Boxes above the graphs indicate between groups total p-value.

**Supplementary Figure 3.** Serum vitamin D [25(OH)D] Levels and Deficiency Status Stratified by Recreational Sun Exposure

Data were presented as bar charts displaying median and interquartile range (IQR) for continuous serum 25(OH)D levels, with median differences and corresponding 95% confidence intervals shown below the graphs. Asterisks denote statistical significance between groups as follows: * = p <0.050, ** = p <0.010, *** = p <0.001, **** = p <0.0001. Dichotomous vitamin D status was presented as stacked bar charts, with frequencies and percentages of participants indicated within the bars. Boxes above the graphs indicate between groups total p-value.

**Supplementary Figure 4.** Serum vitamin D [25(OH)D] Levels and Deficiency Status Stratified by Sun Avoiding Behaviors

Data were presented as bar charts displaying median and interquartile range (IQR) for continuous serum 25(OH)D levels, with median differences and corresponding 95% confidence intervals shown below the graphs. Asterisks denote statistical significance between groups as follows: * = p <0.050, ** = p <0.010, *** = p <0.001, **** = p <0.0001. Dichotomous vitamin D status was presented as stacked bar charts, with frequencies and percentages of participants indicated within the bars. Boxes above the graphs indicate between groups total p-value.

**Supplementary Figure 5.** Association Between Serum vitamin D [25(OH)D] Levels and Deficiency Status with Laboratory Parameters

Data were presented as bar charts displaying median and interquartile range (IQR) for continuous serum 25(OH)D levels, with median differences and corresponding 95% confidence intervals shown below the graphs.

Associations between laboratory parameters and serum 25(OH)D levels were presented as scatter plots with fitted linear regression lines. Correlation coefficients and corresponding *p*-values are indicated within the plots.

