## Supplementary figures and images for "Vitamin D Deficiency among Industrial Workers in Cikarang, Indonesia: Prevalence, Occupational Determinants, and Health Implications"

### Supplementary Figure 1

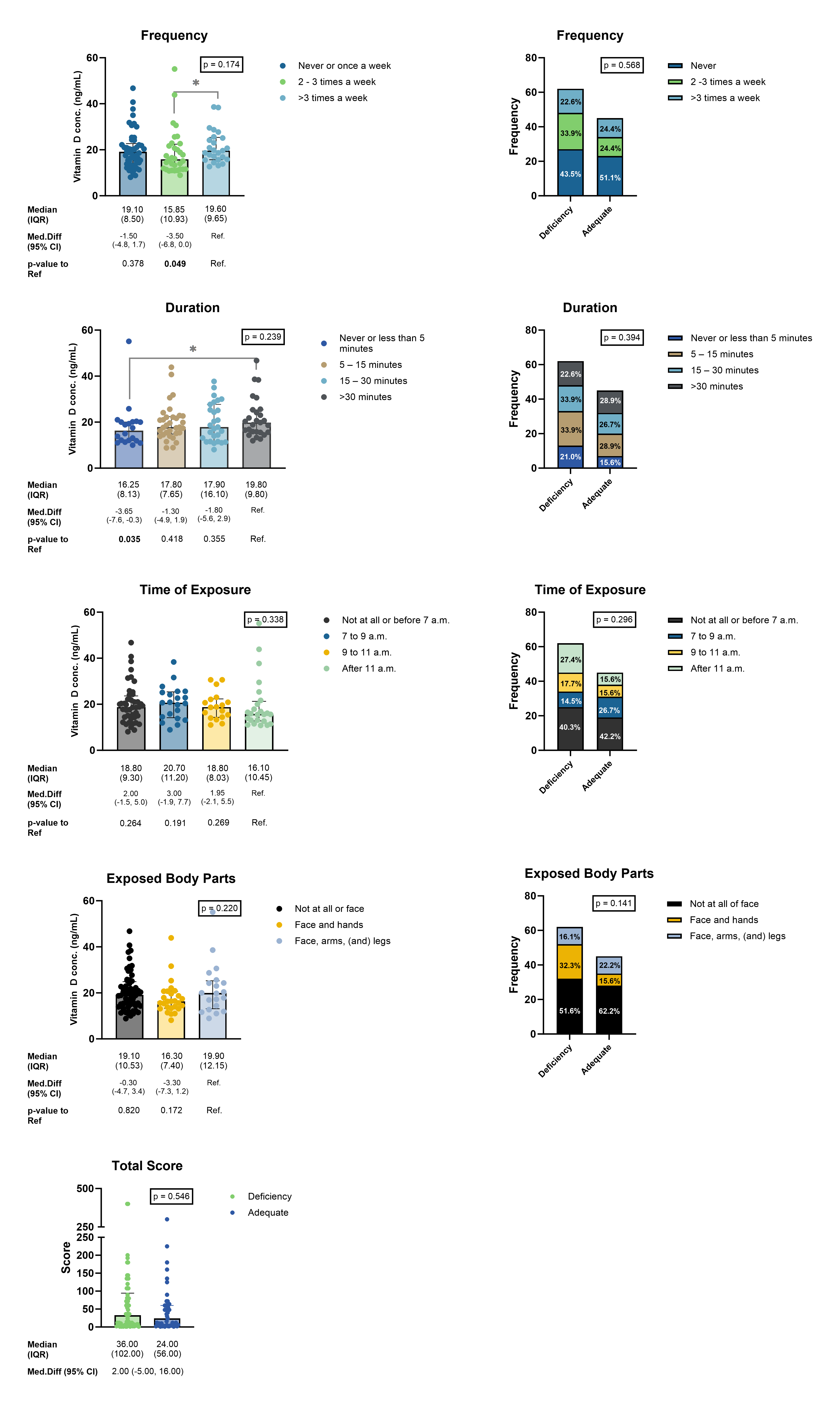

### Supplementary Figure 2

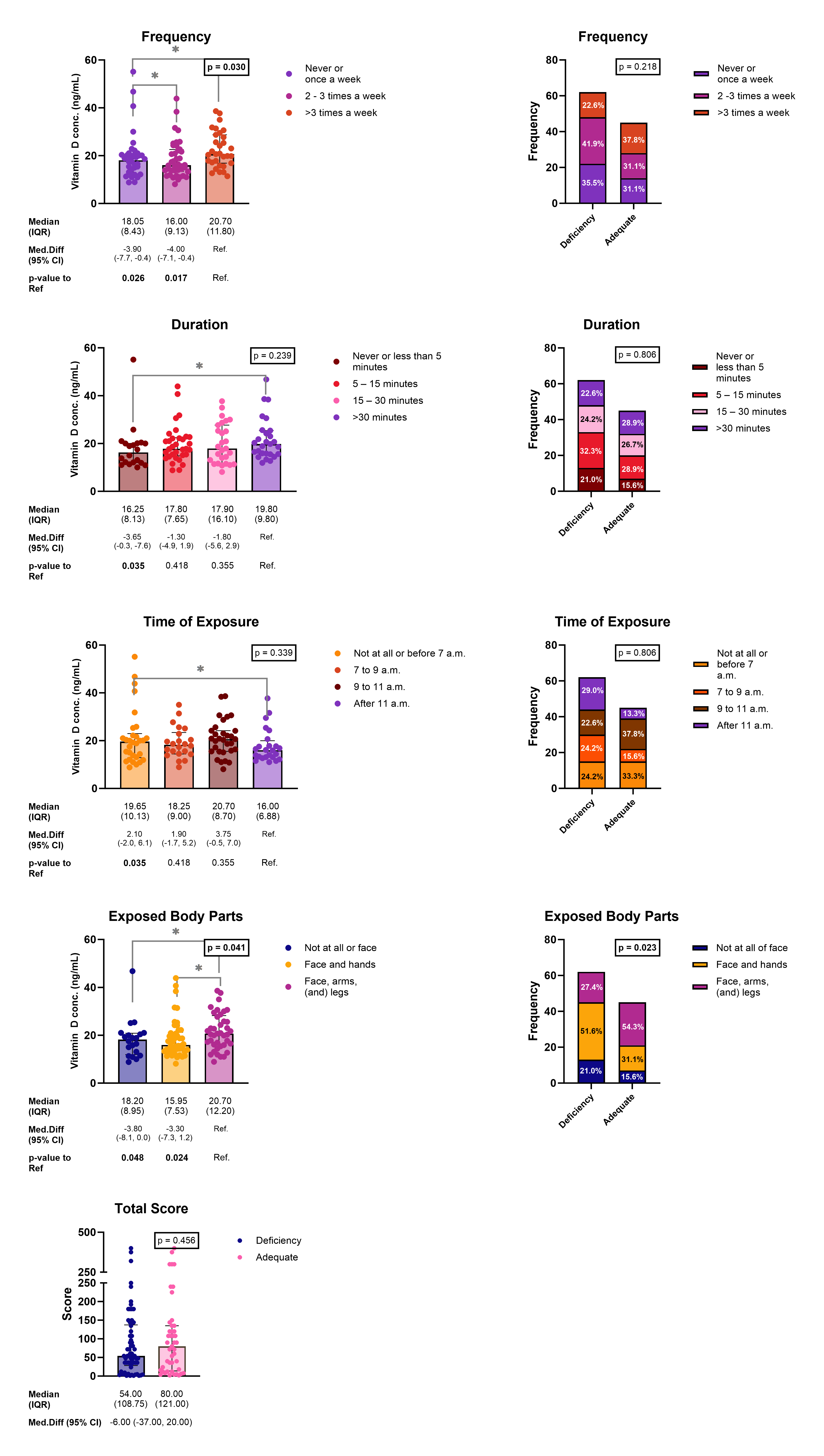

### Supplementary Figure 3

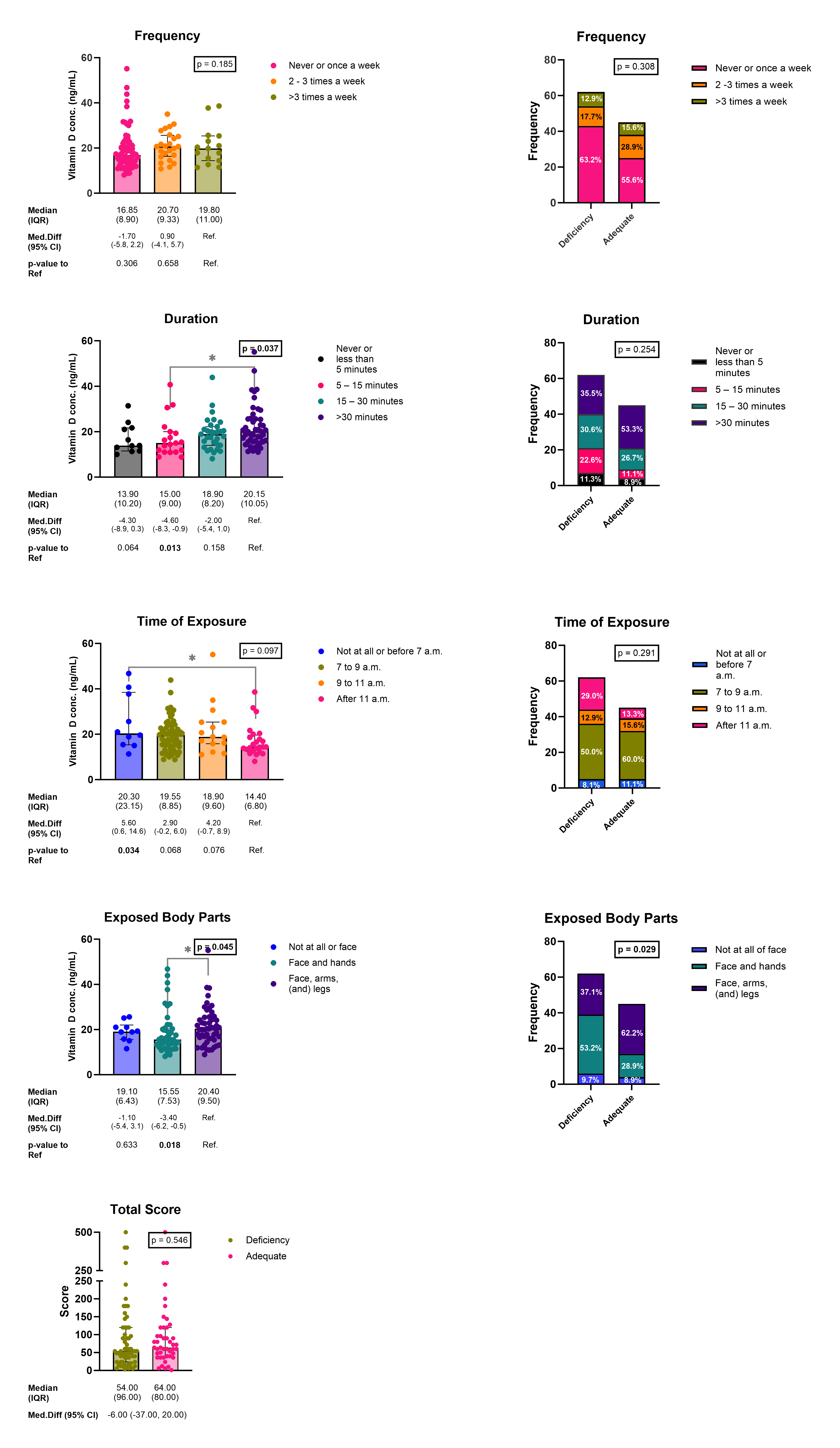

### Supplementary Figure 4

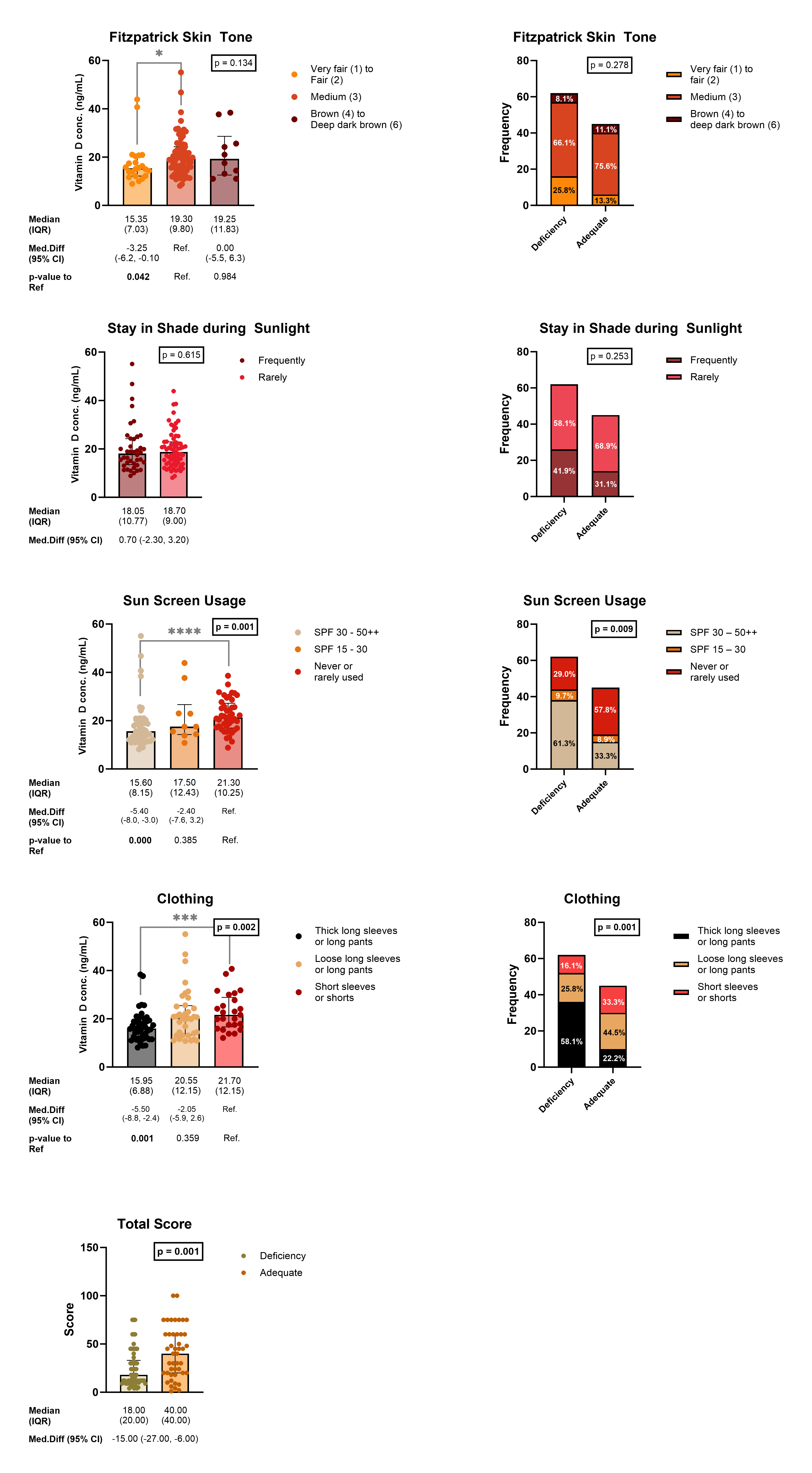

### Supplementary Figure 5

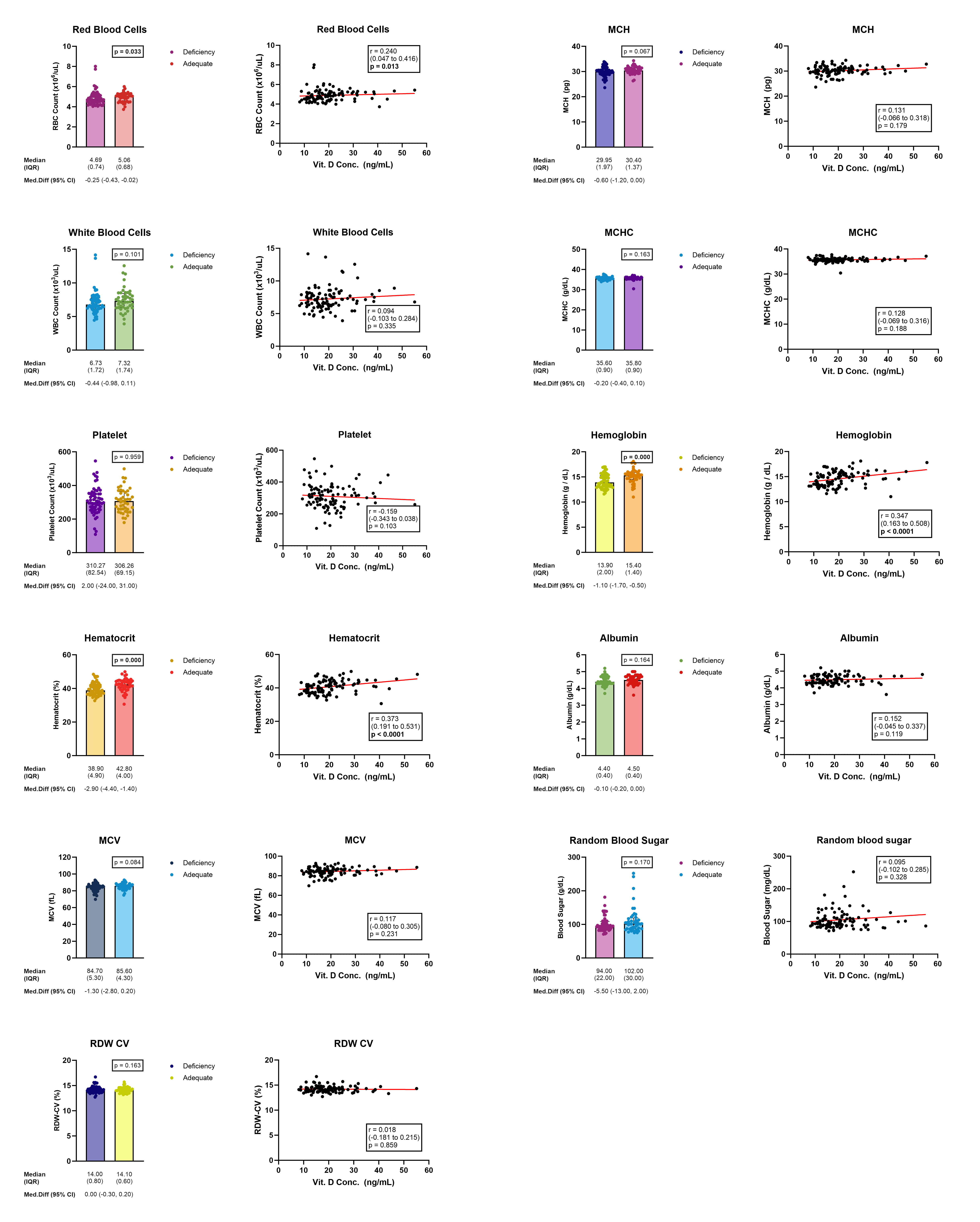
